# Overcoming a diagnostic blindspot: Identifying balanced translocations in Mendelian rare disease cohorts

**DOI:** 10.64898/2026.09.30.26363717

**Authors:** Tejas Jammihal, Kaitlyn Szot, Maninder Kaur, Cecelia Esteves, Shilpa Nadimpalli Kobren, Undiagnosed Diseases Network, Laura Conlin, Ramakrishnan Rajagopalan

## Abstract

**Background:** Short-read genome sequencing generates the signal necessary to detect balanced reciprocal translocations at single-nucleotide resolution, but standard structural variant callers represent them as breakend pairs. Structural variant callers produce thousands of breakend records per genome, the majority of which are false positives arising from segmental duplications, repetitive elements, and reference assembly artifacts. This specificity problem has prevented balanced translocation detection from entering routine genome sequencing workflows. We developed a highly specific pipeline to resolve balanced reciprocal translocations and applied it across five clinically distinct cohorts to characterize the prevalence and diagnostic contribution of balanced reciprocal translocations.

**Methods:** Short-read genome sequencing data from 16,131 individuals across five cohorts were analyzed: healthy participants from the 1000 Genomes Project (n=3,202), probands with structural birth defects and their family members (CHOP BDB cohort; n=2,875, including 1017 probands), probands enrolled in the GREGoR consortium (GREGoR cohort; n=3,574 probands), probands with undiagnosed disease and their family members enrolled in the Undiagnosed Diseases Network (UDN cohort; n=6,071 including 2,027 probands) and neonates who underwent rapid genome sequencing (CHOP Baby Eagle cohort; n=409). A computational pipeline was developed to characterize breakend events from standard structural variant callers to high-confidence balanced translocation calls by exploiting well-characterized properties of true balanced translocation signatures in short-read data. Translocation breakpoints were resolved to single-nucleotide precision and interrogated for disruption of known disease genes or potential regulatory consequence otherwise. A subset of the identified breakpoints was confirmed by Sanger sequencing.

**Results:** Balanced reciprocal translocations were identified in 0.196% of structural birth defect probands (2/1,017), 0.25% of the GREGoR probands (9/3,574), 0.39% of the UDN probands (8/2,027) and 0.24% of neonates who underwent rapid genome sequencing (1/409). Parental carrier frequency in the birth defects cohort was 0.269% (5/1,858), consistent with previously reported estimates in similarly ascertained families, and 0.098% (3/3,040) in the UDN cohort.

All six translocations identified in the birth defects cohort were orthogonally validated using Sanger sequencing, confirming pipeline specificity. Approximately 31% of translocations across the three affected cohorts disrupted a known disease gene at the breakpoint, establishing or refining a molecular diagnosis in five cases previously unresolved. The single translocation identified in the newborn cohort disrupted no disease gene and was classified as a variant of uncertain significance.

## INTRODUCTION

Despite the broad adoption of exome and genome sequencing in research and clinical settings, genetic etiology remains unexplained for a substantial proportion of affected individuals.[1–3] This persistent diagnostic gap may be partially attributable to the putative causal variants beyond single-nucleotide variants and small indels that are missed by exome-focused, routine analytic pipelines.[4–6] Structural variants (SVs) are a major contributor to genetic disorders, and SVs involving copy-number changes (gain or loss of material) are increasingly being detected with high confidence using genome sequencing data. While unbalanced genomic rearrangements are reliably detected by genome sequencing assays (GS), balanced rearrangements such as reciprocal translocations and inversions are largely invisible to routine analysis workflows. Balanced rearrangements can disrupt genes directly at breakpoints, generate aberrant fusion transcripts, separate genes from their cis-regulatory elements, or perturb topologically associated domains (TADs). The developmental consequences of such disruptions are well documented in large cohorts of intellectual disability, autism spectrum disorder, and congenital anomalies.[7, 8]

Conventional cytogenetic karyotyping detects balanced translocations and large inversions at a resolution of approximately 5-10 Mb, missing a large proportion of cryptic events that fall below this threshold.[9, 10] Moreover, karyotype-driven analyses are increasingly impractical given the severe shortage of trained clinical cytogenetics technologists and laboratory directors.[11] Whole-genome sequencing (WGS) offers, in principle, detection of all SV classes at base-pair resolution, and short-read WGS is now routinely applied to large cohorts because of its mature analytical ecosystem. However, balanced rearrangement detection from short-read data remains a major unsolved challenge in practice. Widely used SV callers such as DRAGEN/Manta, DELLY, GRIDSS, and LUMPY routinely emit thousands of breakpoint junctions as breakends without an assigned SV class including hundreds of interchromosomal junctions mimicking a balanced translocation.[12–15] The vast majority of the reported breakends are false positives arising from mismapping in repetitive elements, segmental duplications, and reference-genome artifacts. Among true positives, balanced reciprocal translocations are particularly difficult to disambiguate from sequence insertions, templated insertions, and complex local rearrangements that share overlapping paired-read signatures.[16] Tools such as GATK-SV attempt to detect structural variants comprehensively by integrating variant calls from multiple variant callers and reconstructing complex events from these breakends.[17] However, GATK-SV is currently optimized for use in a specific cloud environment which may be a limitation that prevents wider adoption.[18] The practical consequence is that, even when WGS data are available, balanced translocations are rarely reported, and their population-level contribution is almost certainly underestimated.[19]

To address this gap, we developed a workflow that reconstructs balanced reciprocal translocations from short-read WGS by jointly evaluating reciprocal breakend pairs, explicitly distinguishing balanced exchanges from other variant types. The workflow produces a tractable, high confidence call set of balanced rearrangements per sample, in contrast to the thousands of unfiltered breakends emitted by conventional pipelines. Here we apply this workflow to five different cohorts including 1) The 1000 Genomes Project 2) Children’s Hospital of Philadelphia (CHOP) Birth Defects Biorepository (BDB), 3) GREGoR Consortium, 4) The Undiagnosed Diseases Network (UDN) and 5) the CHOP Baby Eagle newborn genome sequencing datasets. We used these cohorts to (i) validate the workflow by confirming the previously known balanced rearrangements, (ii) estimate the prevalence of balanced reciprocal translocations in these cohorts, and (iii) identify and resolve potentially pathogenic cryptic balanced rearrangements in undiagnosed patients.

## MATERIALS AND METHODS

### 2.1 Cohort Description

**Table 1.** Cohort Description.

|  |  |  |
| --- | --- | --- |
| 1000 Genomes | Healthy participants; reference population cohort | n = 3,202 |
| CHOP BDB cohort | Probands with structural birth defects & family members; 1,017 probands | n = 2,875 |
| CHOP RIMGC cohort | Neonates; rapid genome sequencing cohort | n = 409 |
| GREGoR cohort | Probands enrolled in the GREGoR consortium; 3,574 probands | n = 3,574 |
| UDN cohort | Probands with undiagnosed disease & family members; 2,027 probands | n = 6,071 |

#### 1000 Genomes cohort

The 1000 Genomes Project (1KGP) high-coverage cohort comprises publicly available whole-genome sequencing data from 3,202 individuals (2,504 unrelated individuals plus family members, including 602 trios) spanning 26 populations across five continental groups, as previously published. Structural and copy-number variant call sets (sv.vcf.gz and cnv_sv.vcf.gz) were generated using the DRAGEN v4.4.7 pipeline (using the GRCh38 reference with an integrated pangenome graph), with corresponding CRAM files available for all samples. VCFs were obtained from the publicly hosted AWS S3 bucket (s3://1000genomes-dragen-v4-4-7).[20, 21]

#### CHOP Birth Defects Biorepository cohort

The Children’s Hospital of Philadelphia (CHOP) Birth Defects Biorepository (BDB) cohort is an IRB-approved protocol (#18–015525), allowing access to biological specimens and longitudinal clinical and research data for future studies on birth defects. All participants provided informed consent for the use of their de-identified data and test results in future birth defect research. The cohort comprised 2,875 samples (1,017 probands and 1,858 parents). Structural and copy-number variant call sets (SV.vcf.gz and CNV_SV.vcf.gz) were generated using the DRAGEN v4.4 pipeline (using the GRCh38 reference, hg38-broad-no-alt), with corresponding CRAM files available for all samples.

#### GREGoR Consortium Cohort

The Genomics Research to Elucidate the Genetics of Rare Diseases (GREGoR) Consortium Cohort was established to investigate challenging rare disease cases and evaluate emerging multi-omic technologies for clinical translation. We performed structural variant calling for 3,574 probands using Manta within the Terra cloud environment, while all downstream analyses were executed locally on a high-performance computing cluster. Each candidate variant was subsequently validated via manual inspection of the raw alignments in the corresponding CRAM files.

#### CHOP Baby Eagle Cohort

The CHOP Baby Eagle cohort consists of research-consented neonates and infants referred for genetics consultation at Children’s Hospital of Philadelphia who underwent rapid genome sequencing through CHOP’s Baby Eagle initiative (CHOP IRB #16-013231) (n = 409). Sequencing was performed using the Rapid Targeted Analysis of the Genome for Infants (rTAG-I) test, as previously described.[3] Data were processed using the DRAGEN v4.2.4 pipeline. Structural and copy-number variant call sets (sv.vcf.gz and cnv_sv.vcf.gz) were generated, with corresponding CRAM files available for all samples.

#### Undiagnosed Diseases Network Cohort

The Undiagnosed Diseases Network (UDN) is a National Institutes of Health (NIH) initiative to diagnose patients with rare, long-unexplained conditions through deep phenotyping and genomic analysis.[22] The UDN cohort used here comprises probands with undiagnosed disease and their family members (n = 6,071, including 2,027 probands) enrolled through the UDN. Sequencing data were reprocessed using the DRAGEN v4.4.7 pipeline by the Harvard Medical School Data Coordinating Center (HMS DCC). Structural and copy-number variant call sets (sv.vcf.gz and cnv_sv.vcf.gz), along with corresponding CRAM files, were downloaded from the central UDN data repository (UDN Gateway) for the analysis.

### 2.2 Balanced Translocation Detection

#### BND clustering

Breakend (BND) records in the SV VCF were retained only if they passed the caller’s FILTER, had a SVTYPE of BND, and had a parsable VCFv4.2 ALT allele from which the local and mate breakend orientations were inferred. Both the record’s own chromosome and its mate chromosome were required to lie on primary contigs (chromosomes 1–22, X, Y, and M). Records with an endpoint on an alt, random, decoy, or unplaced contigs were excluded. Records lacking both an EVENT tag and a MATEID were dropped. Breakends were assembled into candidate events in two stages. Each record was first assigned to an initial cluster in one of two ways: EVENT tagged BNDs were grouped by their EVENT value, and untagged BNDs were grouped by their canonical sorted (ID, MATEID) pair so that each junction’s two mates received the same cluster key. Initial clusters sharing a chromosome pair were subsequently merged using a spatial union-find procedure: two clusters were merged if their BNDs were within 1,000 bp of each other on at least one shared chromosome. This two-stage, caller-agnostic approach bridges EVENT-tagged and MATEID-only fragments of the same biological event across both DRAGEN and Manta VCFs. Clusters with fewer than four BND records were discarded, as reciprocal balanced rearrangements require at least two junctions (i.e., four breakends).

#### Difficult-region filtering

Surviving clusters were filtered against a two-tier set of difficult genomic regions. BND records overlapping assembly gaps or centromeres resulted in hard exclusion of the entire cluster. BND records overlapping the ENCODE Blacklist v2 or falling in regions of low 24-mer mappability (k24 Umap track; threshold < 0.7) were annotated with a BLACKLIST_REGION or LOW_MAPPABILITY flag but retained, as breakpoints non-unique to 24-mers are routinely resolvable by 150 bp reads; CRAM-level read evidence was used as the authoritative discriminator for repetitive-region false positives.

#### CRAM-based validation

When a sample CRAM file was available, each BND was annotated with the set of distinct mate chromosomes observed among reads in a 500 bp window around the breakpoint (minimum five supporting reads at MAPQ ≥ 0); split-read inward and outward counts were also recorded. Clusters were then subjected to a cluster-level filter: any cluster in which a BND had reads whose mates mapped to chromosomes not implicated by the cluster’s own call was excluded. This filter exploits the expectation that reads at a true translocation breakpoint should have mates exclusively on the two chromosomes involved in the event; mate evidence on additional chromosomes is characteristic of segmental duplication or repeat-driven false positives. When CRAM files were unavailable (e.g., during cohort-level batch processing), this filter was omitted; the cram_validated output column is set accordingly, and a higher false-positive rate in interchromosomal calls is expected.

#### Topology classification and annotation

Each surviving cluster was classified by two criteria. First, clusters spanning two or more chromosomes were classified as interchromosomal; clusters confined to a single chromosome as intrachromosomal. Second, within each category, clusters were classified as translocations if the maximum span of BND positions on any single chromosome was less than 50 bp, consistent with two junctions at essentially the same genomic coordinate or as insertions if this distance is greater than 50 bp, indicating a donor segment of finite length. The three final event classes were interchromosomal translocation, interchromosomal and intrachromosomal insertions. Putative balanced translocation events were then annotated with gene context and disease associations using the MANE Select transcript set (MANE.GRCh38.v1.5) and the Gene Curation Coalition (GenCC) database, listing genes whose bodies fall within 200 kb of either junction.

### 2.3 Sanger Sequencing Confirmation

Whole blood and saliva sample DNA extractions were performed by CHOP’s Bio Repository Center using QIAsymphony DSP DNA Midi Kit, QIAsymphony DSP DNA Mini Kit and QIAsymphony Sample Prep instrument (QIAGEN, Germany) by standard manufacturer’s protocols. Using the breakpoint location, PCR primers and sequencing primers were generated using Primer3Plus, an automated primer design program. PCR products were tested and then purified using ExoSAP-IT (Thermo Fisher Scientific, United States). The purified PCR product was later sequenced by CHOP’s Center for Applied Genomics (CAG) using the BigDye Terminator Kit v3.1 and ABI 3500 Genetic Analyzer (Applied Biosystems, United States) by standard manufacturer’s protocols. The analysis of the sequenced data was performed using Sequencher Software 5.4.6 (Gene Codes Corporation, United States). Genotypes were verified by manual observation of fluorescent peaks.

## RESULTS

### 3.1 Summary

We applied the balanced translocation detection workflow to whole-genome sequencing data from 16,131 samples across five cohorts: the CHOP Birth Defects Biorepository (BDB; n = 2,875), the NIH Undiagnosed Diseases Network (UDN; n = 6,071), the NHGRI GREGoR Consortium (n = 3,574), CHOP Baby Eagle newborn genome sequencing (n = 409), and the 1000 Genomes Project (n = 3,202). In total we identified 46 unique balanced reciprocal translocations: 6 in BDB, 11 in UDN, 9 in GREGoR, 1 in Baby Eagle and 19 in 1000 Genomes. A t(9;22) junction in one UDN parent (UDN5) corresponded to BCR::ABL1 and was likely somatic, so it is reported separately and excluded from these counts. Across the four disease cohorts, 27 events were identified: eight were diagnostic (five new, three previously established), five were candidates, seven were of uncertain significance, and seven were carried by unaffected relatives.

#### CHOP BDB Cohort

We applied our balanced translocation detection pipeline to whole-genome sequencing data from 2,875 individuals across 1,133 families in the BDB cohort. This population comprised 1,017 probands (534 male, 483 female) and 1,858 parents (848 male, 1,010 female), and 824 families (72.7%) were complete parent–proband trios. The workflow initially returned seven candidate events. After excluding one false positive flagged by the repeats filter, six events across six distinct families remained, carried by seven individuals. All six events were confirmed by read-level validation of the source CRAM files and by Sanger sequencing across the breakpoints.

Two events were found in probands (BDB1, *de novo*; BDB3/BDB4, transmitted from a parent), and the other four were carried by parents only (BDB2, BDB5, BDB6, BDB7), so five of the seven carriers were parents. The proband carried an unbalanced derivative in three of the four parent-only families (BDB2, BDB5, BDB7), and in the fourth (BDB6) the translocation was not transmitted. One event was diagnostic (BDB1, MED13L), one was of uncertain significance (BDB3/BDB4), and four were classified as parental carriers. This corresponds to one event per 479 individuals (0.21%), in line with the estimated general-population frequency of reciprocal translocations (∼1 in 500).

#### GREGoR cohort

We applied the workflow to 3,574 probands (1,964 male, 1,602 female, 8 unknown) in the GREGoR cohort using Manta VCFs only. We identified 9 balanced translocation events, all confirmed by read-level validation of the source CRAM files. For samples with available parental data, running the same analysis showed that two events were maternally inherited (GREGoR4, GREGoR7), giving 9 events in 10 carriers. Five events were *de novo* (GREGoR2, GREGoR3, GREGoR5, GREGoR8, GREGoR9); one was apparently *de novo*, absent in the tested mother with the father not sequenced (GREGoR6); two were maternally inherited (GREGoR4, GREGoR7); and inheritance could not be assessed for one singleton proband with no parental data (GREGoR1).

Four events were diagnostic. Two were previously established diagnoses that disrupt HPRT1 (GREGoR2) and DMD (GREGoR8), and two were new diagnoses in TANC2 (GREGoR3) and ENG (GREGoR6) that are consistent with the patient phenotypes. Three events were candidates (GREGoR1, GREGoR5, GREGoR7) and two were of uncertain significance (GREGoR4, GREGoR9). This corresponds to one event per 397 probands (0.25%).

#### UDN cohort

We applied our balanced translocation detection workflow to the reprocessed whole-genome sequencing data from 6,071 individuals across 2,050 families in the UDN cohort, using DRAGEN structural variant calls. This population comprised 2,027 probands (1,026 male, 999 female), 3,040 parents (1,432 fathers, 1,608 mothers), and 1,004 other relatives (siblings, grandparents, and additional extended family). Of the 2,050 families, 1,382 (67.4%) had a complete parent–proband trio (proband plus both biological parents). Of these, 908 were exact trios and the remaining 474 also included one or more additional sequenced relatives.

The workflow identified 11 balanced translocation events in 13 carriers, and all were confirmed by read-level review of the source CRAM files. Eight events were found in probands, one of which was also carried by an affected father and an affected brother (UDN8–UDN10). The other three were found in unaffected relatives (two mothers, UDN12 and UDN13, and one brother, UDN14). Of the eight proband events, five were *de novo* (UDN1, UDN3, UDN6, UDN7, UDN11), one was paternally inherited (UDN8–UDN10), and two could not be assessed because no parental data were available (UDN2, UDN4, singletons). No unbalanced derivative was detected in the proband for either maternal carrier (UDN12, UDN13), and the event in UDN14 had a homology-mediated junction with an associated deletion.

Three events were diagnostic: one previously established (UDN1, BRSK2) and two new (UDN7, ARID1B; UDN8–UDN10, EXT1). Two events were candidates (UDN2, PCDHA cluster; UDN6, MEIOSIN and RAF1), three were of uncertain significance (UDN3, UDN4, UDN11), and three were carried by unaffected relatives. Five of the eight proband events were known before enrolment (UDN1, UDN2, UDN3, UDN6, UDN11). This corresponds to one event per 552 individuals (0.18%).

Separately, a t(9;22)(q34.12;q11.23) junction (chr9:130,852,816; chr22:23,291,340) in one unaffected father (UDN5) corresponded to BCR::ABL1. It is more consistent with a somatic event (clonal haematopoiesis) than a constitutional translocation, so we excluded it from the counts above.

#### Baby Eagle cohort

In the CHOP Baby Eagle newborn genome sequencing cohort (n = 409), the workflow identified one balanced translocation, t(3;14)(q13.11;q32.33) (BE1; chr3:105,610,628; chr14:106,345,386), in a proband. Inheritance could not be determined. The chr14 breakpoint lies within the IGH locus, with ALCAM and CBLB at or adjacent to the breakpoints, and the event was classified as of uncertain significance. This corresponds to one event per 409 individuals (0.24%).

#### Novel diagnoses

We identified four novel diagnostic variants in the GREGoR (n=2) and the UDN cohorts (n=2) with at least one breakpoint falling within a known disease gene consistent with the proband’s phenotype.

A t(3;9)(q22.3;q34.11) translocation in participant in the proband GREGoR6 was resolved to precise breakpoints at chr3:137131663 and chr9:127839301. The chr9 breakpoint falls in the intron 2 of the ENG affecting all refseq transcripts of the gene. **(Figure 2D)** Heterozygous loss-of-function variants in ENG are an established cause of autosomal dominant hereditary hemorrhagic telangiectasia type 1 (HHT1; MIM #187300), a vascular disorder characterized by mucocutaneous telangiectases, recurrent epistaxis, and arteriovenous malformations. The proband’s reported phenotype -pulmonary arteriovenous malformation (HP:0006548), epistaxis (HP:0000421), and telangiectasia (HP:0001009) is concordant with this diagnosis, making disruption of ENG the definitive molecular explanation for the presentation.

**Figure 1.**
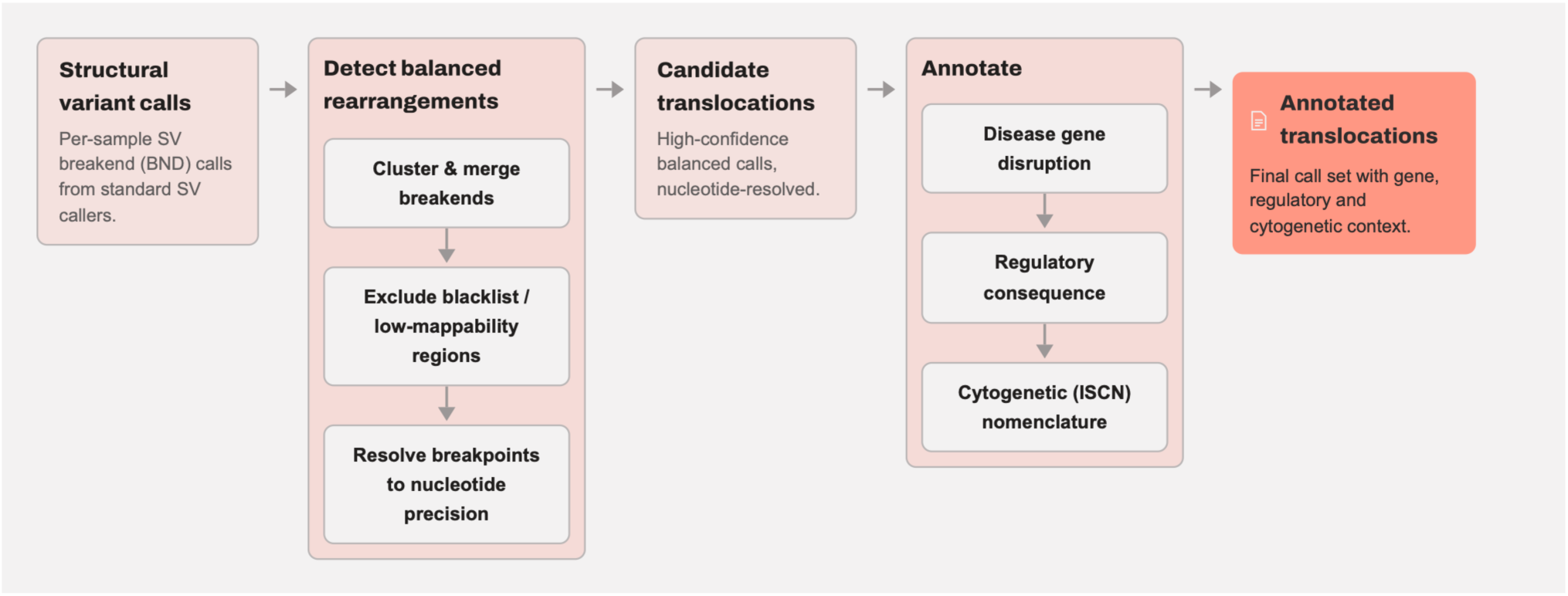
Overview of the bioinformatics pipeline.

**Figure 2.**
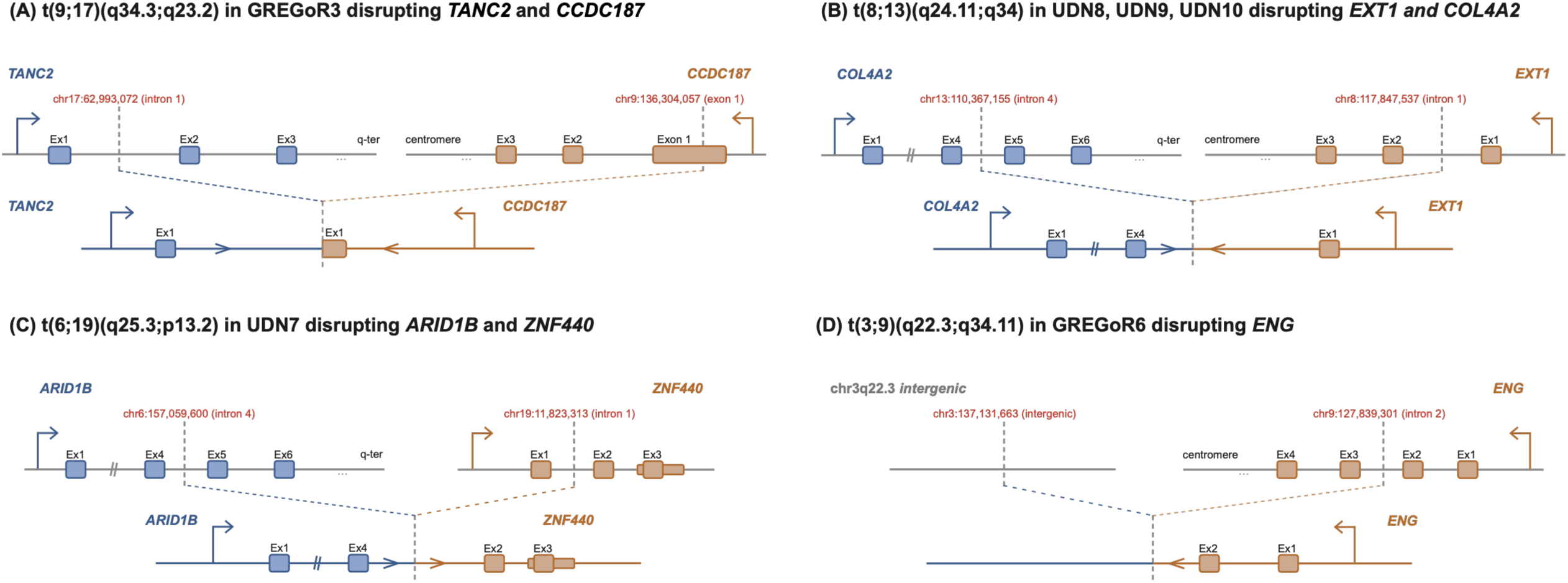
Balanced translocations disrupting disease-associated genes in the GREGoR and UDN cohorts. Schematics of four balanced translocations, showing the two reference chromosomes (top) and the clinically relevant derivative chromosome (bottom). Dashed lines mark the breakpoints, annotated with genomic coordinates and the feature disrupted. Boxes are exons and only exons flanking the breakpoints are shown. Bent arrows indicate transcription start sites and direction of transcription; double slashes indicate omitted sequence. Not drawn to scale. **(A)** t(9;17)(q34.3;q23.2) in GREGoR3 disrupts TANC2 and CCDC187 (neurodevelopmental disorder). **(B)** t(8;13)(q24.11;q34) in a family of three affected individuals (UDN8, UDN9, UDN10) disrupts EXT1 (multiple exostoses) and COL4A2. **(C)** t(6;19)(q25.3;p13.2) in UDN7 disrupts ARID1B (Coffin-Siris syndrome spectrum) and ZNF440. **(D)** t(3;9)(q22.3;q34.11) in GREGoR6 disrupts ENG (hereditary hemorrhagic telangiectasia).

A *de novo* t(9;17)(q34.3;q23.2) translocation in participant GREGoR3 was resolved to precise breakpoints at chr17:62993072 and chr9:136304057. The chr17 breakpoint falls within the intron 1 of TANC2 **(Figure 2A)** known to cause Intellectual developmental disorder with autistic features and language delay, with or without seizures (MIM #618906).[23] The proband is noted to have Global developmental delay (HP:0001263), Frontal bossing (HP:0002007), Cirrohsis (HP:0001394), and Eczematoid dermatitis (HP:0000964). While this proband’s phenotype is sparse, TANC2 is known to be severely constrained for loss-of-function (LOF) variants in the common population (gnomAD v4 LOEUF = 0.17) and LOF variants have been previously reported as pathogenic in several patients.

A *de novo* t(6;19)(q25.3;p13.2) translocation in the UDN7 was resolved to nucleotide resolution and the chr6 breakpoint fell within the intron 4 of the ARID1B (MANE transcript NM_001374828.1) **(Figure 2C)** The proband is noted to have Microcephaly (HP:0000252), Glaucoma (HP:0000501), Retinal dystrophy (HP:0000556), Microdontia (HP:0000691), Delayed speech and language development (HP:0000750), Adrenal insufficiency (HP:0000846), Hypertrichosis (HP:0000998), Seizure (HP:0001250), Hypotonia (HP:0001252), Absent speech (HP:0001344), Asthma (HP:002099), Delayed gross motor development (HP:0002194), Sleep disturbance (HP:0002360), Developmental regression (HP:0002376), Snoring (HP:0025267), and Recurrent ear infections (HP:0410018). Previous genetic workup for the proband included a negative clinical whole exome sequencing which reported a single likely pathogenic variant in RALGAPA1 followed by a negative whole genome sequencing on a research basis. Pathogenic variants in ARID1B cause disease via haploinsufficiency and are classically associated with Coffin-Siris syndrome (MIM #135900). [24] However, ARID1B is also linked to an allelic spectrum of disorders, with over 100 individuals documented in the literature presenting with pathogenic variants outside the classic Coffin-Siris phenotype.[25] The chr19 breakpoint falls within intron 1 of ZNF440 (NM_152357.3). ZNF440 encodes a KRAB-domain zinc-finger transcription factor with no established Mendelian disease association, and we did not consider its disruption further in the interpretation of this case.

Finally, a paternally inherited t(8;13)(q24.11;q34) was identified in a family of 3 affected individuals (UDN8, UDN9, UDN10) with a phenotype of multiple exostoses. The chr8 breakpoint disrupts intron 1 of EXT1 **(Figure 2B)**. Haploinsufficiency is the established disease-causing mechanism for EXT1 and pathogenic variants in EXT1 are known to cause Exostoses, multiple type 1 (MIM #133700).[26] The proband had a significant familial history with the father and the younger brother affected with the same phenotype, and previous negative genetic testing included the panel sequencing of PTPN11 and EXT1/ EXT2.

The chr13 breakpoint falls within intron 4 of COL4A2, N-terminal to the collagenous triple-helical domain in which the majority of characterized pathogenic COL4A2 variants are located. COL4A2 is associated with autosomal dominant brain small vessel disease 2A (BSVD2A; MIM #614483), a cerebrovascular and ocular disorder that most often presents prenatally or in early life with porencephaly, schizencephaly, intracerebral hemorrhage, or retinal abnormalities.[27] Most reported pathogenic COL4A2 variants are missense substitutions within the triple-helical domain thought to act through a dominant-negative mechanism; a loss-of-function splice variant closer in nature to a translocation-mediated disruption has been reported (c.1776+1G>A, intron 24; OMIM #614483.0007), but that variant was inherited from an asymptomatic mother who did not undergo cerebral imaging, illustrating uncertain penetrance for putative loss-of-function COL4A2 alleles. None of the three affected individuals in this family has a reported cerebrovascular, ocular, or structural brain phenotype consistent with BSVD2A, and the exostosis phenotype segregating in this pedigree is fully explained by EXT1 disruption. We therefore consider the COL4A2 breakpoint an incidental finding rather than a contributor to this family’s diagnosis.

#### Confirmation of previously known balanced translocations

Eight participants carried translocations corresponding to molecular diagnoses already established in (1 in BDB, 2 in GREGoR and 5 in UDN), providing a positive control for the pipeline’s ability to resolve diagnostic rearrangements. Four of these corresponded to diagnoses: three previously established (GREGoR2, GREGoR8, UDN1), and one (BDB1) in which the translocation was known but the disrupted gene was first identified here. The other four were known UDN translocations without an established diagnosis (two candidates and two of uncertain significance).

A *de novo* t(10;12)(q24.2;q24.21) translocation in BDB1 was previously known from prior testing, but its exact breakpoints had remained unresolved. Our analysis mapped the precise breakpoints to reveal that the rearrangement directly disrupts MED13L in the intron 4. Heterozygous loss-of-function variants in MED13L are established causes of an autosomal dominant syndrome characterized by intellectual disability and cardiac anomalies (MED13L syndrome; Impaired intellectual development and distinctive facial features with or without cardiac defects; OMIM #616789), making this disruption the definitive molecular diagnosis for the proband’s phenotype.

In GREGoR2, a *de novo* t(X;17)(q26.2;p13.3) translocation was mapped to breakpoints at chr17:1752847 and chrX:134488277, with the chrX breakpoint disrupting HPRT1 the definitive cause of X-linked Lesch-Nyhan syndrome (OMIM #300322), consistent with the proband’s reported phenotype. In GREGoR8, a *de novo* t(X;6)(p21.2;q14.1) translocation (chr6:76851989; chrX:31349387) together with a linked *de novo* deletion at the DMD locus was found to disrupt DMD, the definitive cause of X-linked Duchenne muscular dystrophy (OMIM #310200). In both cases GREGoR records the finding as fully explaining the phenotype, and our breakpoint-level analysis independently recovered the causal gene, confirming these established diagnoses.

For the five UDN cases, the translocations had each already been characterized prior to enrollment, allowing us to assess whether our pipeline could independently recover the known breakpoints and causal loci. In UDN1, a t(11;17)(p15.5;q22) translocation was mapped to breakpoints at chr11:1446810 and chr17:54168880, disrupting BRSK2, the certain cause of a BRSK2-related neurodevelopmental disorder, consistent with the proband’s previously reported balanced translocation between chromosomes 11 and 17. In UDN2, a t(5;7)(q31.3;q34) translocation (chr5:140964614; chr7:140799136) was found to disrupt the PCDHA@ protocadherin cluster, a highly likely candidate locus; the translocation had been identified prior to the participant’s enrollment in the UDN, and whole-genome sequencing here allowed refinement of the breakpoints to base-pair resolution. In UDN3, a *de novo* reciprocal t(X;10)(p11.23;q25.2) translocation (chrX:53864950; chr10:110924649) was originally identified by karyotype, with the normal X chromosome shown to be late-replicating in all cells, consistent with skewed X-inactivation favoring expression of the derivative X. No gene is disrupted at either breakpoint, and the event is of uncertain significance. In UDN6, a *de novo* t(3;19)(p25.2;q13.32) translocation (chr3:12659872; chr19:45753960) disrupting MEIOSIN and RAF1 genes had been previously identified and cytogenetically confirmed.[28] In UDN11, a *de novo* t(7;16)(p22.3;p12.2) translocation (chr7:1651532; chr16:23819173) with no gene at either breakpoint, was likewise a previously established finding of uncertain significance. Across all five UDN cases, our breakpoint-level analysis reproduced the known translocation coordinates, further validating the pipeline’s ability to resolve clinically established rearrangements.

#### Balanced translocations in the 1000 genomes cohort

Across 3,202 individuals screened in the 1000 Genomes cohort, we identified 19 balanced translocation events in 20 carriers, corresponding to one event per 169 individuals (0.59%). HG01762 and HG01763, members of the same family, shared a single t(10;15)(q26.2;q21.1) event. Seven events had a breakpoint within an immune-receptor locus (IGH, IGK, IGL or TRA/TRD; HG01514, HG02396, HG02786, HG04222, NA18610, NA18967, NA19090). Because 1000 Genomes DNA is derived from lymphoblastoid cell lines, these may reflect somatic immune-receptor rearrangement rather than constitutional events.

Five of the 19 events had been previously reported and independently validated by orthogonal methods. Four were identified by Dong et al. (2018), who confirmed each by G-banded karyotype and Sanger sequencing. In HG02260 (t(9;14)(q34.2;q31.1)), the breakpoint disrupted NRXN3. In HG03729 (t(3;17)(q24;p13.3)), the breakpoint disrupted RAP1GAP2, and two flanking cryptic deletions were independently confirmed by qPCR. In NA18612 (t(16;17)(q23.1;q24.2)), the breakpoint disrupted CFDP1, a finding too subtle to resolve by chromosome banding alone and instead confirmed by metaphase FISH. In NA20764 (t(2;19)(p11.2;p13.3)), breakpoints disrupted REEP1 and SHC2. The fifth event, the shared HG01762/HG01763 translocation, was corroborated by an optical mapping cohort (Levy-Sakin et al., 2019), which reported matching breakpoint coordinates at chr10:127,091,700 and chr15:47,100,619. These are consistent with our calls at chr10:127,076,385 and chr15:47,145,012, given the resolution differences expected between short-read and optical mapping approaches. Across all five previously validated events, our breakpoint-level analysis independently recovered coordinates and gene disruptions consistent with these orthogonal methods.

We also manually reviewed long-read alignments for two events, HG01514 and HG04222. HG01514 was supported on manual review but was not called by Sawfish or Sniffles, and HG04222 was flagged as likely noise on review. The remaining 14 events are novel findings not previously reported in the literature, spanning diverse ancestries and expanding the known landscape of balanced structural variation segregating in the general population.

## DISCUSSION

The rapid adoption of molecular technologies has fundamentally altered the clinical genomics landscape, displacing traditional cytogenetics as the primary standard of care. This industry-wide transition has precipitated a critical shortage of trained cytogenetic technologists,[11, 29, 30] eroding the specialized workforce required for legacy testing. Consequently, while cytogenetics maintains a routine footprint in specific domains like prenatal screening, it has lost its place in broader applications.[31, 32] This creates a severe diagnostic blind spot: certain variant classes historically prioritized by cytogenetic workflows, such as reciprocal balanced translocations, are frequently missed by standard genome-wide molecular assays, leading to a profound underappreciation of their true clinical burden. We developed a workflow that leverages short-read sequencing data to identify reciprocal balanced translocations, a variant class traditionally detected through classic cytogenetic techniques.

The central challenge of balanced translocation detection from short-read whole-genome sequencing is not sensitivity but specificity. Standard structural variant callers including Manta/ DRAGEN SV, DELLY, and GRIDSS achieve high sensitivity for interchromosomal junction signal by emitting all discordant read-pair and split-read evidence as breakend (BND) records, without imposing a minimal structural model on the output. The consequence is a per-sample VCF that routinely contains thousands of BND records including hundreds of interchromosomal junctions, the substantial majority of which do not represent true chromosomal rearrangements. The majority are false positives, arising from alignment artifacts in repetitive sequence, segmental duplications, low-complexity regions at putative breakpoints, and the tendency of standard callers to emit spurious reciprocal pairs that mimic the signature of a genuine balanced translocation. This noise burden makes naive application of any standard structural variant caller to clinical cohorts impractical for balanced translocation detection. Manual review of every breakend record at scale is not feasible, and unfiltered call sets cannot support frequency or diagnostic yield estimates with any confidence. The pipeline described here addresses this specificity problem through a series of filters that exploit the well-characterized properties of true balanced translocation signatures in short-read data, reducing the candidate set to a tractable number of high-confidence calls suitable for clinical interpretation.

Several challenges still exist in the comprehensive detection of balanced translocations using short-read sequencing data. Specifically, the identification of balanced rearrangements where repetitive elements flank both sides of the breakpoint is out of scope for this pipeline. This includes any repeat- or segmental duplication-mediated events, such as Robertsonian translocations involving acrocentric chromosomes. Complex rearrangements involving three or more chromosomes, including chromothriptic and chromoplectic events that have been described in constitutional genomic contexts, violate the assumption of reciprocal translocations and may be fragmented into multiple sub-threshold clusters or misclassified as independent pairwise events. The minimum supporting-read thresholds applied throughout the pipeline, five reads for CRAM mate annotation, three per strand for geometry metrics introduce a systematic sensitivity deficit for carriers with low somatic mosaicism and for samples sequenced at coverage below the cohort median. Finally, the workflow provides no phasing information and cannot determine whether two rearrangement breakpoints on the same chromosome are in cis or in trans, a distinction with direct interpretive consequences for compound intrachromosomal rearrangements.

Long-read sequencing platforms, such as PacBio HiFi and Oxford Nanopore Technologies, alongside optical genome mapping from Bionano Genomics, in theory, can achieve markedly superior resolution for balanced rearrangements.[33–36] This advantage maybe especially pronounced at breakpoints nested within repetitive elements where short-read sequencing completely fails.[37] However, automated algorithms to comprehensively identify and annotate these balanced structural variants from long read sequencing data are still largely underdeveloped. Leveraging short-read sequencing to detect these largely unexplored variants is critical as it is the standard of care platform for many first-tier diagnostic tests.

We applied this workflow to 16,131 genomes across five cohorts and identified 46 unique balanced reciprocal translocations. Translocations were identified in 0.20% of structural birth defect probands, 0.25% of GREGoR probands, 0.39% of UDN probands, 0.24% of neonates undergoing rapid genome sequencing, and 0.59% of individuals in the 1000 Genomes Project. The workflow recovered all eight translocations known from prior testing and resolved new molecular diagnoses in five previously unexplained cases: two in GREGoR (ENG, TANC2), two in the UDN (ARID1B, EXT1), and one in the birth defects cohort, where a translocation known from prior cytogenetic testing was shown to disrupt MED13L only once its breakpoints were mapped.

The frequency of balanced translocations was not elevated in the disease cohorts. The 1000 Genomes carrier rate exceeded that of every disease cohort likely reflecting the possibility of cell-line somatic rearrangements. Across the four disease cohorts, 30% (8/27) of translocations disrupted an established disease gene at a breakpoint, compared with 5% (1/19) in the 1000 Genomes Project. This roughly six-fold enrichment suggests that gene disruption at a breakpoint, rather than the presence of a translocation itself, is what distinguishes clinically relevant events from incidental findings.[38, 39] Distinguishing a diagnostic disruption from an incidental one requires base-pair localization of the junction, a resolution that neither microarray nor karyotype can provide. Frequency alone is therefore not an adequate metric for assessing the diagnostic relevance of this variant class.

Contextualizing these frequencies against published population estimates is not straightforward. The widely cited carrier frequency of approximately 1 in 500 derives from newborn karyotyping studies conducted several decades ago, in a clinical environment oriented primarily toward detecting trisomy 21 and its Robertsonian translocation partners.[40, 41] That ascertainment context enriched for Robertsonian rearrangements involving acrocentric chromosomes and captured reciprocal translocations incompletely, particularly those below the resolution of karyotyping (5-10Mb).[42] The classical figure is therefore a poor comparator for reciprocal translocations detected at genome sequencing resolution. The rapid sequencing cohort used here, while not a population sample, offers a more technically matched reference point. A frequency of 0.24% in neonates sequenced on the same platform and with the same pipeline provides a baseline that the historical literature cannot.

The mechanistic basis for pathogenicity in the gene-disrupting cases does not require chromosomal imbalance. A double-stranded break within the coding sequence or regulatory architecture of a dosage-sensitive gene produces a functional consequence equivalent to a loss-of-function point mutation or intragenic deletion, with no net change in copy number.[7, 43, 44] This renders the variant invisible to chromosomal microarray, which is sensitive to copy number gains and losses but blind to balanced rearrangements by design.[42] Standard karyotyping can detect larger reciprocal translocations but cannot resolve breakpoints to gene level and misses cryptic rearrangements entirely.[39, 45] Short-read genome sequencing occupies a different position in this landscape: split reads and discordant read pairs at the breakpoint junction provide single-nucleotide resolution of the fusion sequence, enabling direct interrogation of what gene or regulatory element has been interrupted.[12–14] The pipeline described here makes that resolution accessible at clinical scale.

This positions balanced translocations as a variant class that falls between the detection capabilities of the two platforms most commonly used in clinical genomics. Chromosomal microarray, still a first-tier test for structural birth defects and developmental disorders in many settings, misses them entirely.[42] Karyotyping detects a subset but cannot interpret them at the resolution needed for gene-level diagnosis. As genome sequencing is being widely adopted as a first-tier or early-tier diagnostic test, the inclusion of a high-specificity balanced translocation detection step in the analytical pipeline represents a tractable addition that recovers a category of pathogenic variation that would otherwise go unrecognized.[46–48]

The distinction between *de novo* and inherited translocations remains relevant to clinical interpretation. *De novo* balanced translocations carry a well-established higher prior probability of pathogenicity compared to those transmitted from a phenotypically normal parent.[49] However, an inherited translocation that disrupts a disease gene is not automatically benign. Variable expressivity and incomplete penetrance can make a carrier parent appear unaffected, and careful phenotypic evaluation of transmitting parents is warranted when a gene-disrupting translocation is identified in a proband.[7] The elevated translocation frequency observed in the parental cohort at 0.25% most likely reflects ascertainment that these parents came to attention because they had a child with a structural anomaly, and a proportion will have transmitted an unbalanced derivative chromosome rather than the balanced form. That ascertainment effect should be considered before interpreting the parental frequency as evidence of enrichment.

Several limitations apply to these findings. The affected cohorts were clinically ascertained rather than population-recruited, and a formally matched unaffected control group sequenced and analyzed with the same pipeline is not available. The absolute numbers of balanced translocations in each cohort are small, which limits the precision of frequency estimates and makes statistical comparison between cohorts unreliable. Pipeline sensitivity, the proportion of true balanced translocations in these genomes cannot be fully characterized without orthogonal validation at scale using long-read sequencing or optical genome mapping. Causal inference for individual gene-disrupting translocations rests on breakpoint evidence and phenotypic correlation, and functional validation was not uniformly available. These findings should therefore be interpreted as a demonstration of the pipeline’s utility and an initial estimate of the diagnostic contribution of this variant class, with larger prospective studies needed to refine both.

## Data Availability

The balanced translocation events identified in this study, including breakpoint coordinates and affected genes, are provided in the manuscript and its tables. The analysis pipeline will be made available on GitHub. Individual-level patient sequencing data cannot be made publicly available owing to participant privacy protections and the terms of the respective consent and data-use agreements. Access to underlying genomic data is governed by the individual cohorts: 1000 Genomes Project data are publicly available as described in the Methods; Undiagnosed Diseases Network data are available through the UDN Gateway and dbGaP to approved researchers; GREGoR Consortium data are available under the consortium's data-use agreement; and CHOP Birth Defects Biorepository and Baby Eagle data are available from the corresponding author upon reasonable request, subject to institutional and IRB approval.

## ACKNOWLEDGEMENTS

This work was supported by the National Human Genome Research Institute of the National Institutes of Health under award number R01HG013355 to R.R. The authors thank the patients and families whose participation made this study possible. We gratefully acknowledge the CHOP Undiagnosed Diseases Program, CHOP Omics & Big Data Initiative, and the Arcus research data platform at Children’s Hospital of Philadelphia for their contributions to this study. The Undiagnosed Diseases Network is supported by the National Institute of Neurological Disorders and Stroke of the National Institutes of Health (U2CNS132415). The GREGoR Consortium is funded by the National Human Genome Research Institute of the National Institutes of Health, through the following grants: U01HG011758, U01HG011755, U01HG011745, U01HG011762, U01HG011744, and U24HG011746. The content is solely the responsibility of the authors and does not necessarily represent the official views of the National Institutes of Health.

